# Impact of Prolonged Obesity Across the Lifecourse on Myocardial Stress and Injury in Older Age: Birth Cohort Study

**DOI:** 10.64898/2026.10.08.26365055

**Authors:** William Johnson, Tom Norris, Alun D. Hughes, Mark Hamer, Steven Bell, G. David Batty

## Abstract

**Background:** The consequences of prolonged exposure to obesity on biomarkers of myocardial stress and injury remain unclear. We examined associations of duration and cumulative severity of obesity from childhood to later adulthood with concentrations of N-terminal pro-B-type natriuretic peptide (NT-proBNP), high-sensitivity cardiac troponin I (hs-cTnI), and growth differentiation factor-15 (GDF-15) at age 62 years.

**Methods:** The sample comprised 4,901 participants in the 1958 National Child Development Study who had measurements of NT-proBNP, hs-cTnI, or GDF-15 at age 62 years and at least four BMI assessments between 7 and 62 years of age. Individual BMI trajectories were modelled, from which total duration of obesity and mean excess BMI (area between the trajectory and the age-specific obesity threshold divided by duration) were derived. Associations of ever versus never having been obese, obesity duration, and mean excess BMI with biomarker concentrations were examined using general linear regression models with progressive adjustment for early-life characteristics; contemporaneous cardiometabolic factors; and health behaviours, socioeconomic position, and psychosocial factors. Nonlinear associations of obesity duration were estimated using restricted cubic splines and subsequently approximated using linear splines.

**Results:** Overall, 26.3% of participants had ever experienced obesity. There was no evidence of an association between ever versus never having been obese and NT-proBNP. Among men, ever having been obese was associated with 11.00% (95% CI, 4.62–17.37) higher hs-cTnI in the fully adjusted model, whereas there was no evidence of an association among women. Ever having been obese was associated with higher GDF-15, although the association attenuated substantially after adjustment for contemporaneous cardiometabolic factors. Longer obesity duration was associated with higher concentrations of all three biomarkers in fully adjusted models, but the pattern differed by biomarker: hs-cTnI increased approximately linearly with duration, whereas NT-proBNP and GDF-15 remained relatively stable up to approximately 25 years of obesity before increasing with longer duration. In fully adjusted models, each additional year of obesity beyond 25 years was associated with 1.80% (95% CI, 0.54–3.06) higher NT-proBNP and 0.81% (95% CI, 0.25–1.36) higher GDF-15, while each additional year across the full range of obesity duration was associated with 0.41% (95% CI, 0.07–0.76) higher hs-cTnI. Among participants who had ever experienced obesity, each 1 kg/m² higher mean excess BMI was associated with 4.02% (95% CI, 1.79–6.24) higher NT-proBNP and 2.00% (95% CI, 0.49–3.51) higher hs-cTnI in fully adjusted models; there was no evidence of an association with GDF-15.

**Conclusions:** Longer duration of obesity was associated with higher concentrations of biomarkers reflecting myocardial stress, injury, and cellular stress at age 62 years, with associations particularly evident after approximately 25 years of obesity for NT-proBNP and GDF-15. These findings suggest that duration of obesity may provide clinically relevant information beyond current obesity status, supporting its consideration as an additional dimension of cumulative obesity burden in cardiovascular risk assessment.

## Introduction

Cardiovascular disease (CVD) remains the leading cause of death globally and a major contributor to morbidity. In 2019, an estimated 523 million people were living with CVD and 18.6 million deaths were attributable to CVD worldwide [1]. Beyond clinically overt disease, cardiovascular pathology can develop progressively over the life course, with subclinical myocardial injury and haemodynamic stress detectable before the onset of clinically apparent disease [2]. Understanding the determinants of these early pathological changes may help identify factors contributing to cardiovascular damage before clinical disease develops. Circulating biomarkers provide sensitive measures of distinct aspects of subclinical cardiovascular pathology, including myocardial wall stress, myocardial injury, and cellular stress. N-terminal pro-B-type natriuretic peptide (NT-proBNP) is released in response to myocardial wall stress and is an established biomarker of cardiac dysfunction and heart failure [3]. High-sensitivity cardiac troponin I (hs-cTnI) provides a sensitive measure of myocardial injury, with low-level circulating concentrations detectable in individuals without clinically overt CVD [4 5]. Growth differentiation factor-15 (GDF-15) is a stress-responsive cytokine that is upregulated in response to cellular stress and has been associated with CVD and adverse cardiovascular outcomes [6 7]. Although GDF-15 is not specific to myocardial stress or injury, its associations with CVD and adverse outcomes provide an opportunity to examine whether cumulative obesity exposure is also associated with broader stress-related cardiovascular processes.

The prevalence of obesity has increased substantially worldwide, with more than 1 billion people now living with obesity [8]. Obesity is an established risk factor for CVD, with excess adiposity contributing to cardiovascular pathophysiology through metabolic, haemodynamic, and inflammatory pathways [9]. Previous studies have generally reported lower NT-proBNP concentrations among individuals with higher BMI or obesity, although the association appears to vary by sex and other characteristics [10–12]. In contrast, higher BMI has been associated with higher concentrations of hs-cTnI and GDF-15, consistent with greater subclinical myocardial injury and cellular stress [13 14]. Measures of obesity at a single time point do not, however, capture the cumulative burden of excess adiposity. Individuals with the same obesity status at a given age may have experienced very different durations and magnitudes of exposure. Longer duration and greater magnitude of obesity may therefore provide additional information about cumulative exposure and its potential relationship with subclinical cardiovascular pathology. Previous studies have examined associations between obesity duration and cardiovascular outcomes, with longer duration generally associated with greater cardiovascular risk [15–18]. However, evidence on the relationship between cumulative obesity exposure and subclinical myocardial stress and injury remains limited.

One small cross-sectional study of 33 patients with severe obesity found that NT-proBNP concentrations were correlated with duration of obesity (ρ = 0.339, P < 0.004), although the paper does not report how the duration of obesity was determined [19]. For hs-cTnI, evidence from longitudinal studies suggests that longer-term obesity exposure is associated with greater subclinical myocardial injury. In the Trøndelag Health Study, participants with stable obesity, compared with those with stable normal weight across four BMI assessments spanning approximately 35 years, had higher hs-cTnI concentrations at the final sweep (median age 68.7 years) [20]. In a related analysis of the same cohort, each 100 excess BMI-years was associated with a 15.6% (95% CI 13.0–18.2) higher hs-cTnI concentration [21]. Similarly, in the Atherosclerosis Risk in Communities (ARIC) study, each additional 10 years of obesity and each 100 excess BMI-years were associated with greater odds of elevated high-sensitivity cardiac troponin T (hs-cTnT), another marker of subclinical myocardial injury [22]. However, these longitudinal studies have relied on a relatively small number of BMI assessments beginning in adulthood and have not characterised obesity exposure from childhood through later adulthood in terms of duration, age at onset, and cumulative severity. For GDF-15, evidence linking long-term obesity exposure with circulating concentrations is more limited. In a longitudinal study of 496 adults with obesity but without diabetes, changes in GDF-15 over 4 years were positively associated with changes in body weight, BMI, and waist-to-hip ratio [23]. More recently, in the Santiago Longitudinal Study of 205 participants, those with persistent obesity from childhood to age 29 years had higher GDF-15 concentrations in young adulthood than those who maintained a healthy BMI from birth to age 29 years [24]. However, evidence remains limited on whether the duration, age at onset, and cumulative severity of obesity are associated with GDF-15 concentrations in later adulthood.

Using repeated measures of BMI from the 1958 National Child Development Study (NCDS), the aim of this study was to investigate the associations of duration of obesity from age 7 to 62 years with concentrations of NT-proBNP, hs-cTnI, and GDF-15 at age 62 years. Because obesity tends to persist once established [25], duration of obesity is very strongly inversely correlated with age at obesity onset; therefore, associations with obesity duration also provide information about the timing of obesity onset. Among individuals who had ever been obese, we also examined associations of cumulative severity of obesity with the biomarkers.

## Methods

### Study

The 1958 NCDS is a prospective birth cohort study comprising 18,558 participants, including 17,638 individuals born during one week in March 1958 in England, Scotland, and Wales and 920 immigrants born during the same week who were incorporated into the cohort during childhood [26].

### Ethics and Informed Consent

Ethical approval for data collection in the 1958 NCDS was obtained from the relevant National Health Service Research Ethics Committees, including the London–Central Research Ethics Committee (reference 12/LO/2010). Ethical approval for the age-62 biomedical sweep was provided by the London–Fulham Research Ethics Committee (reference 18/LO/2034). Ethical and consent procedures reflected the requirements applicable at the time of each sweep. Informed consent, including parental consent during childhood, was obtained in accordance with the procedures applicable to each sweep. The present study involved secondary analysis of anonymised NCDS data and did not involve direct contact with participants. All data used in the present study are freely available to download via the UK Data Service (https://ukdataservice.ac.uk/).

### Sample

A total of 8,405 study members participated in the age 62-year sweep, of whom 4,933 had data on one or more of the outcomes (i.e., NT-proBNP, hs-cTnI, and GDF-15). We excluded 32 participants with fewer than four serial BMI measurements because a minimum of four observations was considered necessary to reliably estimate individual BMI trajectories across the life course using the multilevel spline model (see Exposures section). The final analytical sample therefore comprised 4,901 participants (2,434 men and 2,467 women).

### Outcomes

At the age 62-year sweep non-fasting venous blood samples were collected from participants who provided consent for blood sampling and analysis. A 6-mL K2-EDTA tube was collected and transported to the University of Bristol for storage. Following completion of fieldwork, stored samples were analysed at the BHF Glasgow Cardiovascular Research Centre, University of Glasgow. NT-proBNP was measured using the Roche proBNP II assay on a cobas e801 analyser, with a manufacturer analytical detection range of 5–35,000 pg/mL and a limit of quantitation of 50 pg/mL. hs-cTnI was measured using a chemiluminescent microparticle immunoassay (Abbott hsTnI, Architect i1000 analyser), with a manufacturer analytical detection range of 1.1–50,000 pg/mL and a limit of quantitation of 1.3 pg/mL. GDF-15 was measured using the Roche GDF-15 electrochemiluminescence immunoassay on a cobas e801 analyser, with a manufacturer analytical detection range of 400–20,000 pg/mL and a limit of quantitation of 400 pg/mL. Manufacturer quality-control samples were run during the assays. The 1958 NCDS documentation reports no outliers across the batch of samples for these biomarkers.

For descriptive purposes, biomarker concentrations were categorised using clinically or previously reported thresholds. NT-proBNP concentrations were categorised as <125, 125–<400, and ≥400 pg/mL [27]. hs-cTnI concentrations were categorised according to sex-specific thresholds, with concentrations below 34.2 pg/mL in men and 15.6 pg/mL in women classified as below the threshold, and concentrations at or above these values classified as above the threshold [28]. GDF-15 concentrations were categorised as <1,200, 1,200–1,800, and >1,800 pg/mL, consistent with previously reported GDF-15 categories [29]. These categories were used solely to describe the distribution of biomarker concentrations in the study population and were not used to define clinical diagnoses.

### Exposures

BMI (kg/m²) was derived from height and weight obtained at ages 7, 11, 16, 23, 33, 42, 44, 50, 55, and 62 years; height and weight were self-reported at ages 23, 42, 50, and 55 years, whereas measurements at the other ages were objectively assessed. In the analytical sample, 42,199 BMI observations were available, with 81% of participants having eight or more serial BMI observations.

To derive life course obesity traits (e.g., obesity duration), we modelled individual BMI trajectories from 7-62 years of age within a multilevel framework (measurement occasion at level 1 and individuals at level 2). Chronological age (centered at 7 years) was fitted as a restricted cubic spline, with seven knots at default locations based on Harrell’s recommended quantiles [30]. The model constant and the six age spline terms were allowed to have random effects at level two, with an unstructured variance–covariance matrix, thereby allowing each participant to have their own trajectory. Sex was included as a main effect and as interactions with the six age spline terms. Further, the level one variance (i.e., error) was allowed to differ according to whether the data were measured or self-reported.

Using the multilevel model, annual fitted BMI values from 7 to 62 years of age were obtained for each individual. These fitted values were converted to Z-scores using the International Obesity Task Force (IOTF) reference [31]. Obesity was defined as a Z-score >2.288 in males and >2.192 in females, corresponding to the IOTF BMI cut-off of 30 kg/m² at 18 years of age. The primary exposure, “total duration of obesity”, was calculated as the length of time, in years, that a person’s BMI Z-score exceeded the obesity threshold. “Age at first obesity” was derived, representing the age when BMI Z-score first crossed upwards through the obesity threshold. The “number of times obese” was calculated as the number of times an individual’s BMI Z-score crossed upwards through the obesity threshold. Finally, we used the composite trapezoid rule to derive a cumulative obesity severity variable, represented in Figure 1 by the Area Under the Curve (AUC) and above the obesity threshold. Mean excess BMI was calculated as the AUC divided by the total duration of obesity, representing the mean amount by which BMI exceeded the obesity threshold during periods of obesity.

**Figure 1.**
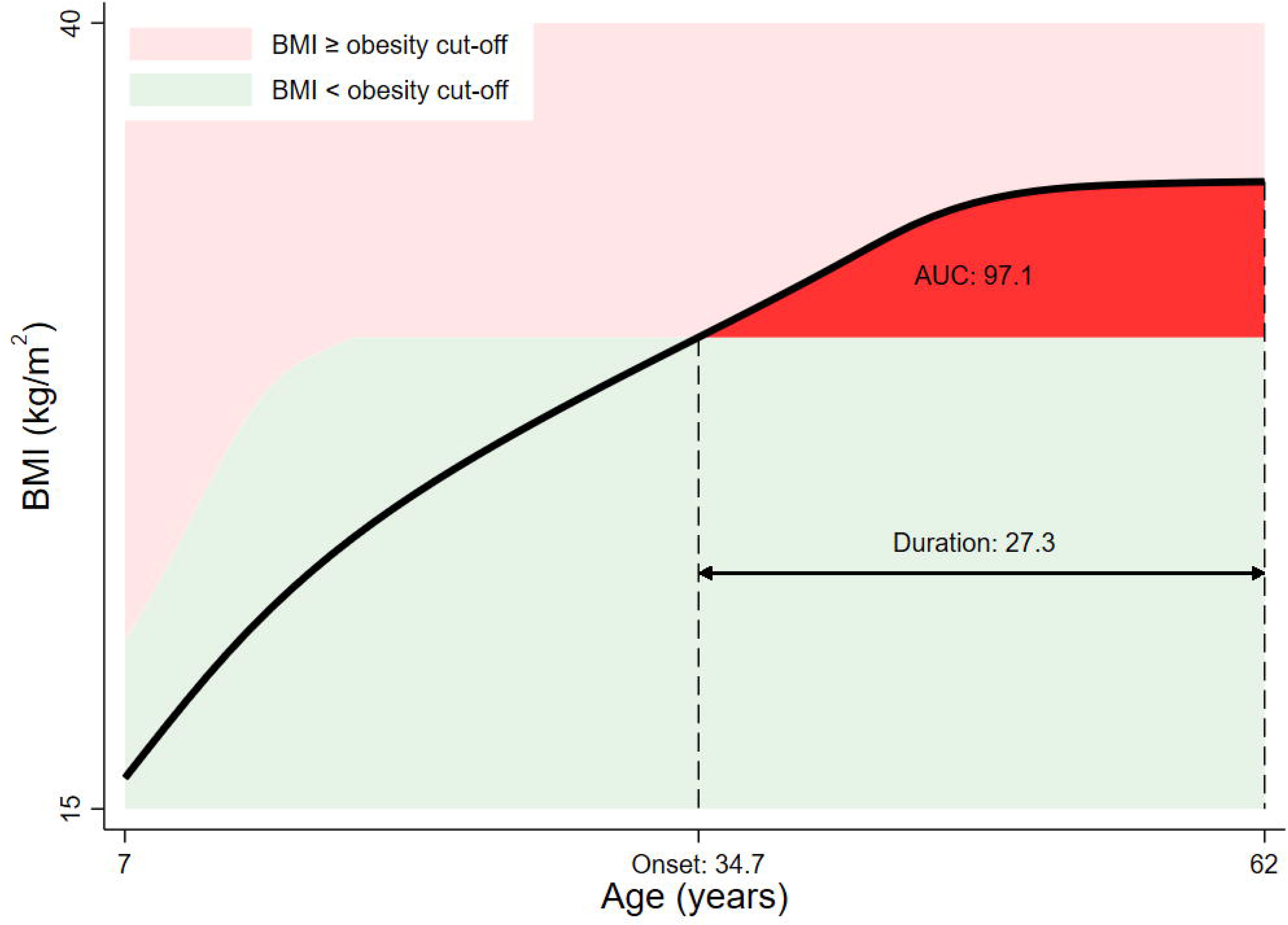
Example obesity traits (onset, duration, and severity (i.e., AUC but above obesity cut-off)) for one participant.

### Covariate set 1. Early life characteristics

In addition to cohort-member sex and ethnicity (white British and other), birth weight was recorded at the birth survey by the attending midwife, with information also obtained from medical records. Father’s occupation, measured when the participant was 11 years old, was classified according to the Registrar General’s Social Class (I professional, II managerial and technical, IIIN skilled non-manual, IIIM skilled manual, IV partly skilled, and V unskilled) [32]. At age 11 years, general cognitive ability was assessed using a standardised General Ability Test administered in schools as part of the 1958 NCDS [33]. The test consisted of 80 items, equally divided between verbal and non-verbal questions. For each item, children selected one of five possible responses, with one point awarded for each correct answer. Scores therefore ranged from 0 to 80, with higher scores representing better cognitive performance. At age 7 years, behavioural and social adjustment was assessed using the teacher-completed Bristol Social Adjustment Guide (BSAG) [34 35]. Teachers indicated the behavioural descriptions that best characterised each child, with responses covering a range of behavioural and emotional characteristics that were classified into 12 behavioural syndromes. An overall BSAG score was derived from the individual items, with higher scores indicating greater behavioural maladjustment.

### Covariate set 2. Contemporaneous cardiometabolic factors

At age 62 years, systolic blood pressure was measured by a trained healthcare professional using an Omron HEM-907 automated monitor, with two seated measurements taken one minute apart; their mean was used in analyses. Non-fasting venous blood samples were collected during the health visit. High-sensitivity C-reactive protein (CRP), total cholesterol and HDL-C were measured using enzymatic/immunoturbidimetric assays, while HbA1c was measured using ion-exchange high-performance liquid chromatography. LDL-C was derived from the available lipid measurements.

### Covariate set 3. Health behaviours, socioeconomic position, and psychosocial factors

At age 62 years, health behaviours and educational attainment were assessed using self-reported questionnaire data. Alcohol consumption was categorised as abstainer, within recommended limits, or exceeding recommended limits based on reported drinking. Smoking status was classified as never smoker, ex-smoker, occasional smoker, or daily smoker. Regular exercise was assessed by self-report and categorised as yes or no. Highest educational qualification was derived from reported qualification histories and recategorised into four groups: none; low (GCSE D–E or Scottish Standard Grade/Nat 5 [CS2–5]); medium (GCSE A–C/Good O-Levels, A-Level or AS-Level, ≥2 A-Levels, or Scottish Higher); and high (Diploma, PGCE or nursing diploma, degree-level qualification [e.g., BA/BSc or equivalent professional qualification], or higher degree [MA/MSc/PhD]). At age 62 years, mental health was assessed using the five-item Mental Health Inventory (MHI-5), derived from the SF-36 health survey [36 37]. Participants reported how frequently during the previous four weeks they had experienced five emotional states: nervousness, feeling downhearted or blue, feeling so down that nothing could cheer them up, calmness and peacefulness, and happiness. Responses were combined to generate a score ranging from 0 to 100, with higher scores indicating better mental health. Mental wellbeing was assessed using the seven-item Shortened Warwick-Edinburgh Mental Well-Being Scale (SWEMWBS) [38]. Participants reported how often during the previous two weeks they had experienced seven positively worded statements relating to mental wellbeing and psychological functioning, using a five-point response scale from 1 (“none of the time”) to 5 (“all of the time”). Item scores were combined and transformed using the recommended SWEMWBS scoring procedure, with higher scores indicating greater mental wellbeing. Cognitive function was assessed using immediate and delayed word-list recall and animal naming. For immediate recall, participants were asked to recall as many as possible of 10 words presented verbally. After a delay, participants had two minutes to recall as many words as possible from the original list. For animal naming, participants were asked to name as many different animals as possible within one minute.

### Statistical analysis

Descriptive statistics were used to summarise participant characteristics, exposures, covariates, and outcome biomarkers, with continuous variables presented as means (SD) or medians (IQR), as appropriate, and categorical variables as frequencies and percentages. The distribution of total duration of obesity was examined using a histogram.

As shown in Table 1, some variables had missing data. Multiple imputation by chained equations (MICE) was used to impute missing values under a missing-at-random (MAR) assumption, following the approach described by Sterne et al [39]. MICE uses fully conditional specification, whereby missing values are imputed iteratively using a sequence of univariate imputation models [40]. General linear regression was used for continuous variables, with variables log-transformed before imputation where appropriate because of skewness; binary logistic regression was used for binary variables; and multinomial logistic regression was used for categorical variables. Where perfect prediction occurred during the imputation of categorical variables, augmented regression was used to address this issue. The imputation model included the three study outcomes, all covariates, and the binary indicator of ever versus never having been obese. Other life-course obesity variables (age at first obesity onset, total duration of obesity, obesity severity [area under the curve; AUC], mean excess BMI, and number of periods of obesity) were not included in the imputation model because these variables were structurally missing for participants who had never been obese. Fifty imputed datasets were generated, and subsequent regression models used Rubin’s rules to pool estimates across the 50 datasets.

**Table 1.** Description of the study sample.

|  |  |  | Missing<br>N (%) |
| --- | --- | --- | --- |
| **Outcomes** |  |  |  |
| NT-proBNP (pg/mL) | Median (IQR) | 64.7 (36.6, 113.0) | 48 (1.0) |
| <125 | N (%) | 3821 (78.0) |  |
| 125 – <400 | N (%) | 928 (18.9) |  |
| ≥400 | N (%) | 104 (2.1) |  |
| hs-cTnI (pg/mL) | Median (IQR) | 3.2 (2.4, 4.6) | 78 (1.6) |
| Below sex-specific threshold | N (%) | 4752 (98.5) |  |
| ≥34.2 (men); ≥15.6 (women) | N (%) | 71 (1.5) |  |
| GDF-15 (pg/mL) | Median (IQR) | 1013 (796, 1346) | 22 (0.5) |
| <1,200 | N (%) | 3257 (66.8) |  |
| 1,200 – 1,800 | N (%) | 1042 (21.4) |  |
| >1,800 | N (%) | 580 (11.9) |  |
| **Obesity traits** |  |  |  |
| Never obese | N (%) | 3611 (73.7) |  |
| Ever obese | N (%) | 1290 (26.3) |  |
| Age first onset (years) | Median (IQR) | 42.0 (35.6, 46.0) |  |
| Total duration (years) | Median (IQR) | 19.5 (12.9, 26.3) |  |
| Severity (AUC) | Median (IQR) | 31.8 (8.1, 88.8) |  |
| Mean excess BMI (AUC/duration) | Median (IQR) | 1.6 (0.6, 3.5) |  |
| Number of periods |  |  |  |
| 1 | N (%) | 1255 (97.3) |  |
| 2 | N (%) | 35 (2.7) |  |
| Correlation (age first onset – total duration) | Spearman's ρ | -0.96 |  |
| Correlation (total duration – mean excess BMI) | Spearman's ρ | 0.87 |  |
| Correlation (age first onset – mean excess BMI) | Spearman's ρ | -0.81 |  |
| **Covariate set 1. Early life characteristics** |  |  |  |
| Sex |  |  |  |
| Male | N (%) | 2434 (49.7) |  |
| Female | N (%) | 2467 (50.3) |  |
| Ethnicity |  |  | 4 (0.1) |
| White British | N (%) | 4805 (98.1) |  |
| Other | N (%) | 92 (1.9) |  |
| Birth weight (kg) | Mean (SD) | 3.35 (0.50) |  |
| Father's occupational class at age 11 years |  |  | 720 (14.7) |
| I (professional) | N (%) | 249 (6.0) |  |
| II (managerial and technical) | N (%) | 954 (22.8) |  |
| IIIN (skilled non-manual) | N (%) | 505 (12.1) |  |
| IIIM (skilled manual) | N (%) | 1654 (39.6) |  |
| IV (partly skilled) | N (%) | 554 (13.3) |  |
| V (unskilled) | N (%) | 265 (6.3) |  |
| General Ability Test at age 11 years | Mean (SD) | 47.7 (14.8) | 585 (11.9) |
| Bristol Social Adjustment Guide at age 7 years | Median (IQR) | 4 (1, 10) | 518 (10.6) |
| **Covariate set 2. Contemporaneous cardiometabolic factors** |  |  |  |
| Systolic blood pressure (mmHg) | Mean (SD) | 132.9 (17.0) | 45 (0.9) |
| CRP (mg/L) | Median (IQR) | 1.2 (0.6, 3.0) | 908 (18.5) |
| HbA1c (mmol/mol) | Median (IQR) | 40 (38, 43) | 137 (2.8) |
| LDL-C (mmol/L) | Mean (SD) | 3.2 (1.0) | 915 (18.7) |
| HDL-C (mmol/L) | Mean (SD) | 1.6 (0.5) | 123 (2.5) |
| **Covariate set 3. Health behaviours, socioeconomic position, and psychosocial factors** |  |  |  |
| Alcohol consumption |  |  |  |
| Abstainer | N (%) | 1438 (29.3) |  |
| Within limits (≤14 units/week) | N (%) | 2346 (47.9) |  |
| Exceeds limits (>14 units/week) | N (%) | 1117 (22.8) |  |
| Smoking |  |  | 1 (0.02) |
| Never | N (%) | 2645 (54.0) |  |
| Ex | N (%) | 1780 (36.3) |  |
| Occasionally | N (%) | 63 (1.3) |  |
| Every day | N (%) | 412 (8.4) |  |
| Regular exercise |  |  | 9 (0.2) |
| Yes | N (%) | 4025 (82.3) |  |
| No | N (%) | 867 (17.7) |  |
| Qualifications |  |  | 293 (6.0) |
| No | N (%) | 489 (10.6) |  |
| Low | N (%) | 1244 (27.0) |  |
| Medium | N (%) | 1670 (36.2) |  |
| High | N (%) | 1205 (26.2) |  |
| Mental Health Inventory (MHI-5) | Mean (SD) | 64.5 (7.6) | 319 (6.5) |
| Warwick-Edinburgh Mental Well-Being Scale (shortened version) | Mean (SD) | 26.5 (4.3) | 374 (7.6) |
| Immediate word-list recall | Mean (SD) | 6.3 (1.5) | 59 (1.2) |
| Delayed word-list recall | Mean (SD) | 5.0 (1.8) | 59 (1.2) |
| Animal naming | Mean (SD) | 23.7 (6.5) | 60 (1.2) |

General linear regression models were used to estimate differences in the three outcomes between participants who had ever versus never been obese. All outcomes were transformed using 100ln (y), such that regression coefficients can be interpreted as symmetric percentage differences. Four sets of models were fitted: (1) unadjusted; (2) adjusted for covariate set 1 (early-life characteristics); (3) additionally adjusted for covariate set 2 (contemporaneous cardiometabolic factors); and (4) additionally adjusted for covariate set 3 (health behaviours, socioeconomic position, and psychosocial factors). Effect modification by sex was assessed by including an interaction term between sex and the ever-versus-never obesity indicator. There was consistent evidence of effect modification by sex for hs-cTnI (P-values for interaction <0.05); therefore, results for this outcome are presented separately for men and women. Equivalent regression models were fitted to examine associations between mean excess BMI and the three outcomes among participants who had ever been obese. We explored non-linear associations using restricted cubic splines but found no evidence of non-linearity.

We next examined the association between total duration of obesity and the three outcomes among participants who were ever obese. To account for potential non-linearity, restricted cubic spline models with four knots placed at the 5th, 35th, 65th, and 95th percentiles of the exposure distribution were fitted. All outcomes were log-transformed. Initial models were unadjusted for covariates, and predicted mean curves with corresponding 95% confidence intervals (CIs) were plotted across the observed range of obesity duration. Predicted values were back transformed to the original outcome scale and therefore represent geometric means. To examine whether associations differed by sex, interaction terms between sex and each spline term were added to the models, and a joint Wald test of the three interaction terms was performed. There was no evidence of effect modification by sex (joint Wald test, P>0.05). Models were then adjusted for mean excess BMI to examine the association of obesity duration independent of the average severity of obesity during periods of obesity. The unadjusted and mean excess BMI-adjusted curves were presented together in a single plot. Finally, we examined the association between obesity duration and the three outcomes after progressive adjustment for the prespecified covariate sets. The unadjusted model and three progressively adjusted models were presented together in a single plot.

While the association of total obesity duration with hs-cTnI was approximately linear, the associations with NT-proBNP and GDF-15 were non-linear. Between 0.055 years (i.e., the lowest observed obesity duration) and approximately 25 years of obesity duration, there was little evidence of an association, with the curves remaining approximately flat. In contrast, between approximately 25 and 55 years, both outcomes increased with increasing duration of obesity. For hs-cTnI, we fitted general linear regression models with a single linear term for obesity duration. For NT-proBNP and GDF-15, we fitted linear spline models with a single knot at 25 years to estimate separate slopes for obesity duration below 25 years and 25 years and above. We formally tested whether the slope changed after 25 years; there was consistent evidence of a change in slope for both NT-proBNP and GDF-15. As in the previous analyses, all outcomes were transformed using 100ln (y), and four sets of models with progressively greater levels of covariate adjustment were fitted. Effect modification by sex was assessed by including interaction terms between sex and obesity duration; for the spline models, interactions with both spline terms were included and assessed using a joint Wald test. There was no evidence of effect modification by sex.

All statistical analyses were performed using Stata version 19 (StataCorp LLC, College Station, TX, USA). The runmlwin command was used to fit the multilevel models [41].

### Code availability

Available from the first author upon request.

## Results

Descriptive statistics are shown in Table 1. Median concentrations were 64.7 pg/mL (IQR 36.6–113.0) for NT-proBNP, 3.2 pg/mL (IQR 2.4–4.6) for hs-cTnI, and 1,013 pg/mL (IQR 796–1,346) for GDF-15. Most participants had NT-proBNP <125 pg/mL (78.0%) and hs-cTnI below the sex-specific threshold (98.5%), while 66.8% had GDF-15 <1,200 pg/mL. Overall, 26.3% of participants had ever experienced obesity. Among these participants, median age at first onset was 42.0 years (IQR 35.6–46.0), median total duration of obesity was 19.5 years (IQR 12.9–26.3), and median mean excess BMI was 1.6 kg/m² (IQR 0.6–3.5). Most participants with obesity had one period of obesity (97.3%), while 2.7% experienced two periods. Age at first obesity onset was strongly inversely correlated with total duration of obesity (Spearman’s ρ=−0.96), while total duration was strongly positively correlated with mean excess BMI (ρ=0.87). The distribution of total duration of obesity is shown in Supplementary Figure 1.

The associations of ever versus never having obesity with the three outcomes are shown in Table 2. Model 1 was unadjusted; Model 2 additionally adjusted for early-life characteristics; Model 3 further adjusted for contemporaneous cardiometabolic factors; and Model 4 additionally adjusted for health behaviours, socioeconomic position, and psychosocial factors. Across all models, there was no evidence of an association with NT-proBNP. The association between ever having obesity and hs-cTnI differed by sex. Among men, ever having obesity was associated with higher hs-cTnI, with only minimal attenuation across models with increasing levels of covariate adjustment. In Model 4, hs-cTnI was 11.00% higher (95% CI 4.62–17.37) among participants who had ever been obese than among those who had never been obese. Among women, there was no evidence of an association between ever having obesity and hs-cTnI. Ever having obesity was associated with higher GDF-15 across all models, although the association attenuated substantially after adjustment for contemporaneous cardiometabolic factors, from 18.51% (95% CI 15.76– 21.25) in Model 2 to 5.46% (95% CI 2.65–8.27) in Model 3.

**Table 2.**
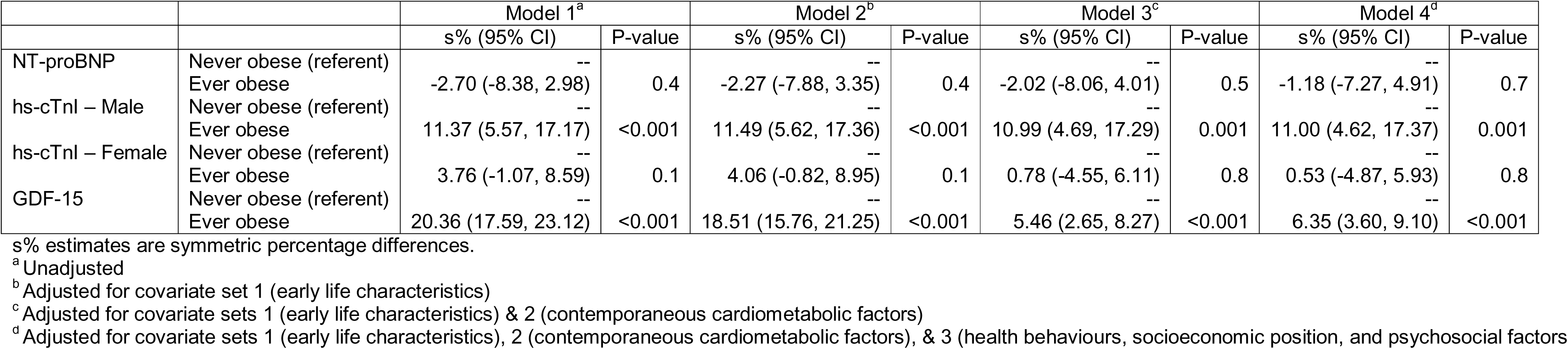
The associations of ever versus never obese with NT-proBNP, hs-cTnI, and GDF-15.

|  |  | Model 1<sup>a</sup> |  | Model 2<sup>b</sup> |  | Model 3<sup>c</sup> |  | Model 4<sup>d</sup> |  |
| --- | --- | --- | --- | --- | --- | --- | --- | --- | --- |
|  |  | s% (95% CI) | P-value | s% (95% CI) | P-value | s% (95% CI) | P-value | s% (95% CI) | P-value |
| NT-proBNP | Never obese (referent) | -- |  | -- |  | -- |  | -- |  |
|  | Ever obese | -2.70 (-8.38, 2.98) | 0.4 | -2.27 (-7.88, 3.35) | 0.4 | -2.02 (-8.06, 4.01) | 0.5 | -1.18 (-7.27, 4.91) | 0.7 |
| hs-cTnI – Male | Never obese (referent) | -- |  | -- |  | -- |  | -- |  |
|  | Ever obese | 11.37 (5.57, 17.17) | <0.001 | 11.49 (5.62, 17.36) | <0.001 | 10.99 (4.69, 17.29) | 0.001 | 11.00 (4.62, 17.37) | 0.001 |
| hs-cTnI – Female | Never obese (referent) | -- |  | -- |  | -- |  | -- |  |
|  | Ever obese | 3.76 (-1.07, 8.59) | 0.1 | 4.06 (-0.82, 8.95) | 0.1 | 0.78 (-4.55, 6.11) | 0.8 | 0.53 (-4.87, 5.93) | 0.8 |
| GDF-15 | Never obese (referent) | -- |  | -- |  | -- |  | -- |  |
|  | Ever obese | 20.36 (17.59, 23.12) | <0.001 | 18.51 (15.76, 21.25) | <0.001 | 5.46 (2.65, 8.27) | <0.001 | 6.35 (3.60, 9.10) | <0.001 |

s% estimates are symmetric percentage differences.
<sup>a</sup> Unadjusted
<sup>b</sup> Adjusted for covariate set 1 (early life characteristics)
<sup>c</sup> Adjusted for covariate sets 1 (early life characteristics) & 2 (contemporaneous cardiometabolic factors)
<sup>d</sup> Adjusted for covariate sets 1 (early life characteristics), 2 (contemporaneous cardiometabolic factors), & 3 (health behaviours, socioeconomic position, and psychosocial factors)

Figures 2–4 show the unadjusted associations between total obesity duration and the three outcomes. For NT-proBNP and GDF-15, concentrations remained relatively stable up to approximately 25 years of obesity duration, after which they increased with longer duration. Linear spline regression analyses supported these patterns (Table 3). Across all models, there was no evidence of an association between obesity duration and NT-proBNP or GDF-15 when duration was <25 years, whereas beyond 25 years, each additional year was consistently associated with higher concentrations of both biomarkers. In Model 4, each additional year beyond 25 years of obesity duration was associated with a 1.80% (95% CI 0.54–3.06) higher NT-proBNP and a 0.81% (95% CI 0.25–1.36) higher GDF-15. In contrast, the association between obesity duration and hs-cTnI was approximately linear (Figure 3), with positive estimates across all models, although the association was not statistically significant in the unadjusted model (Table 3). In Model 4, each additional year of obesity duration was associated with a 0.41% (95% CI 0.07–0.76) higher hs-cTnI.

**Figure 2.**
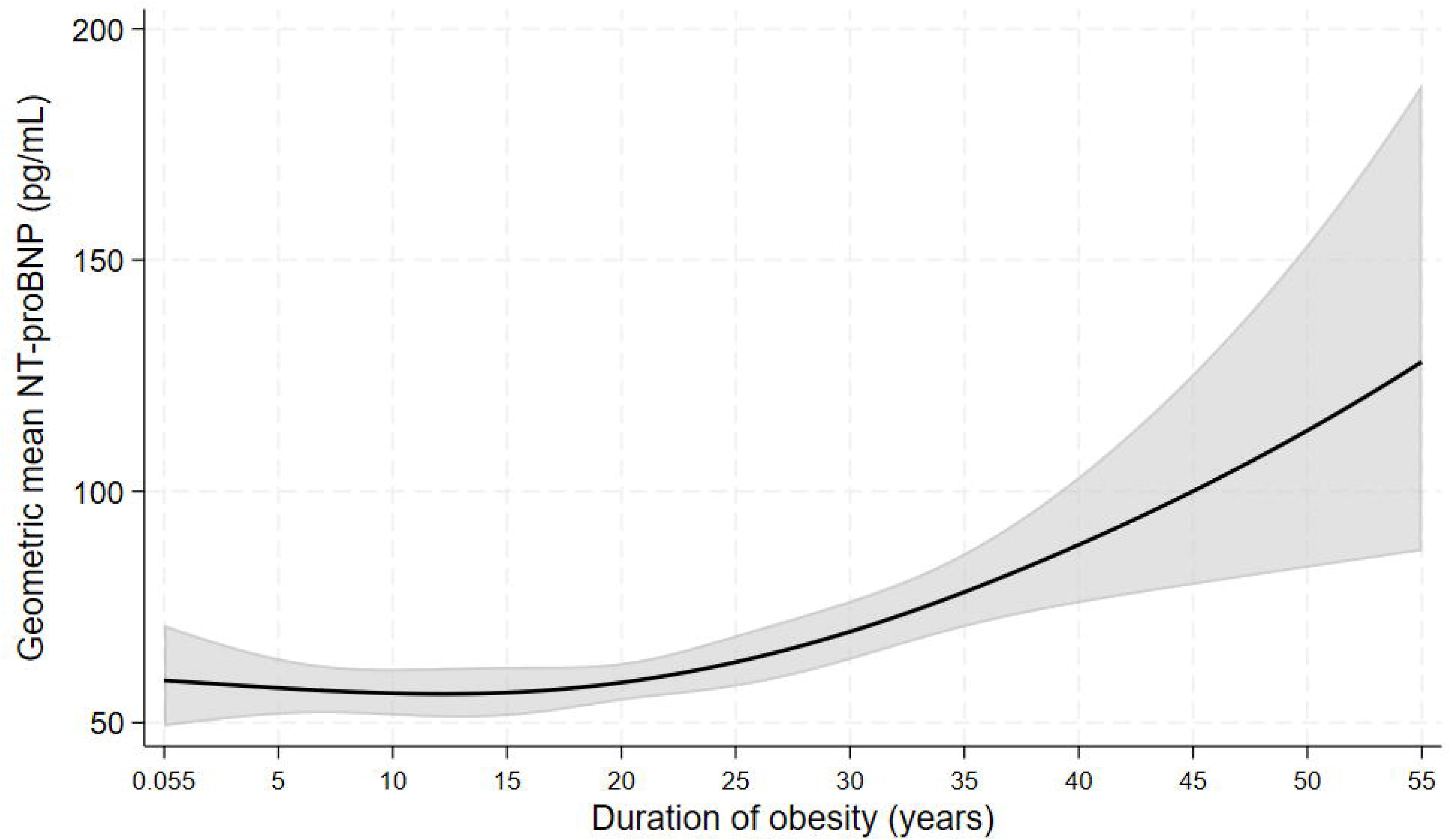
The association of obesity duration with NT-proBNP (pg/mL) among participants who were ever obese, unadjusted for covariates in. **Table 1**

**Figure 3.**
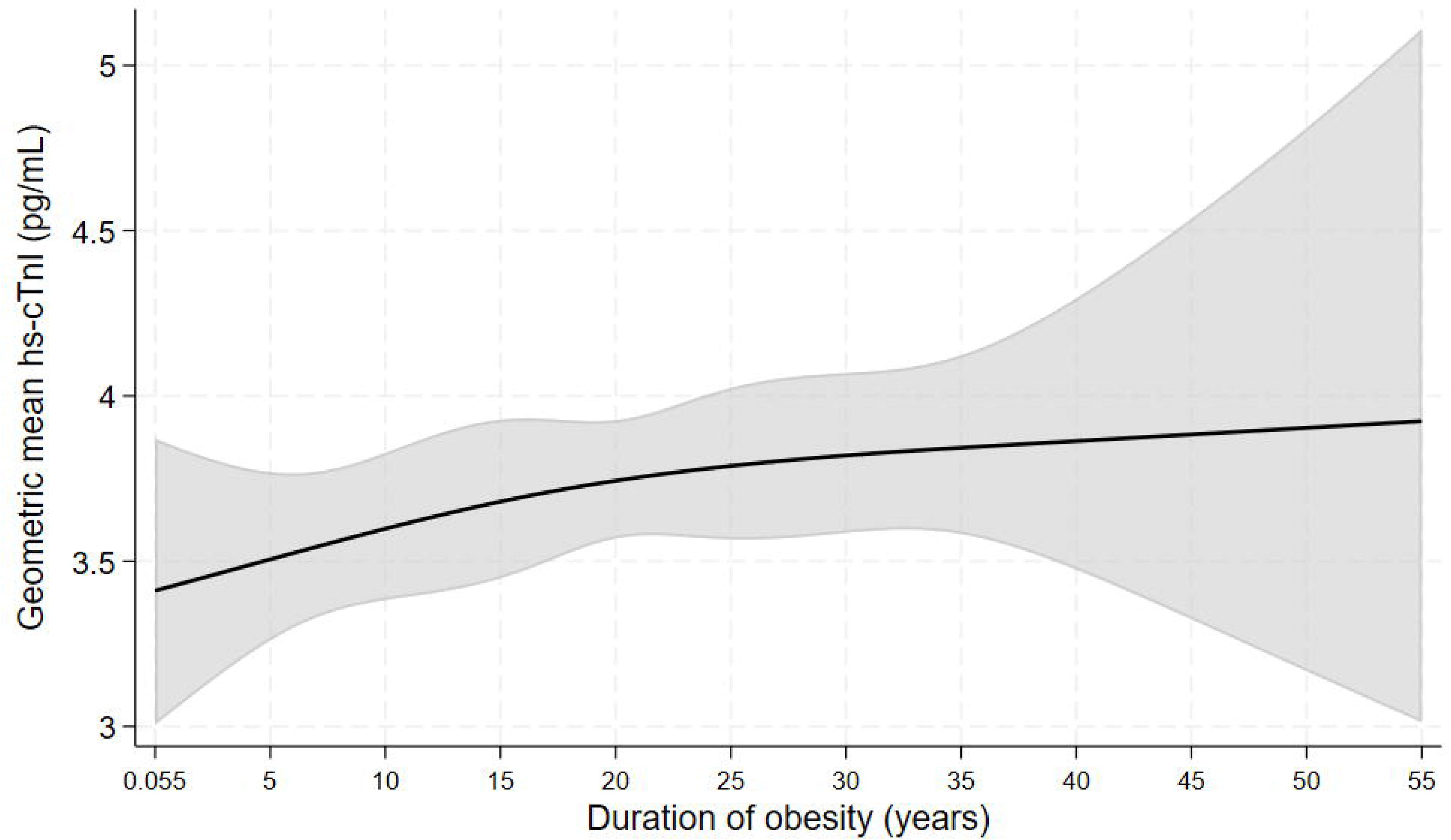
The association of obesity duration with hs-cTnI (pg/mL) among participants who were ever obese, unadjusted for covariates in. **Table 1**

**Figure 4.**
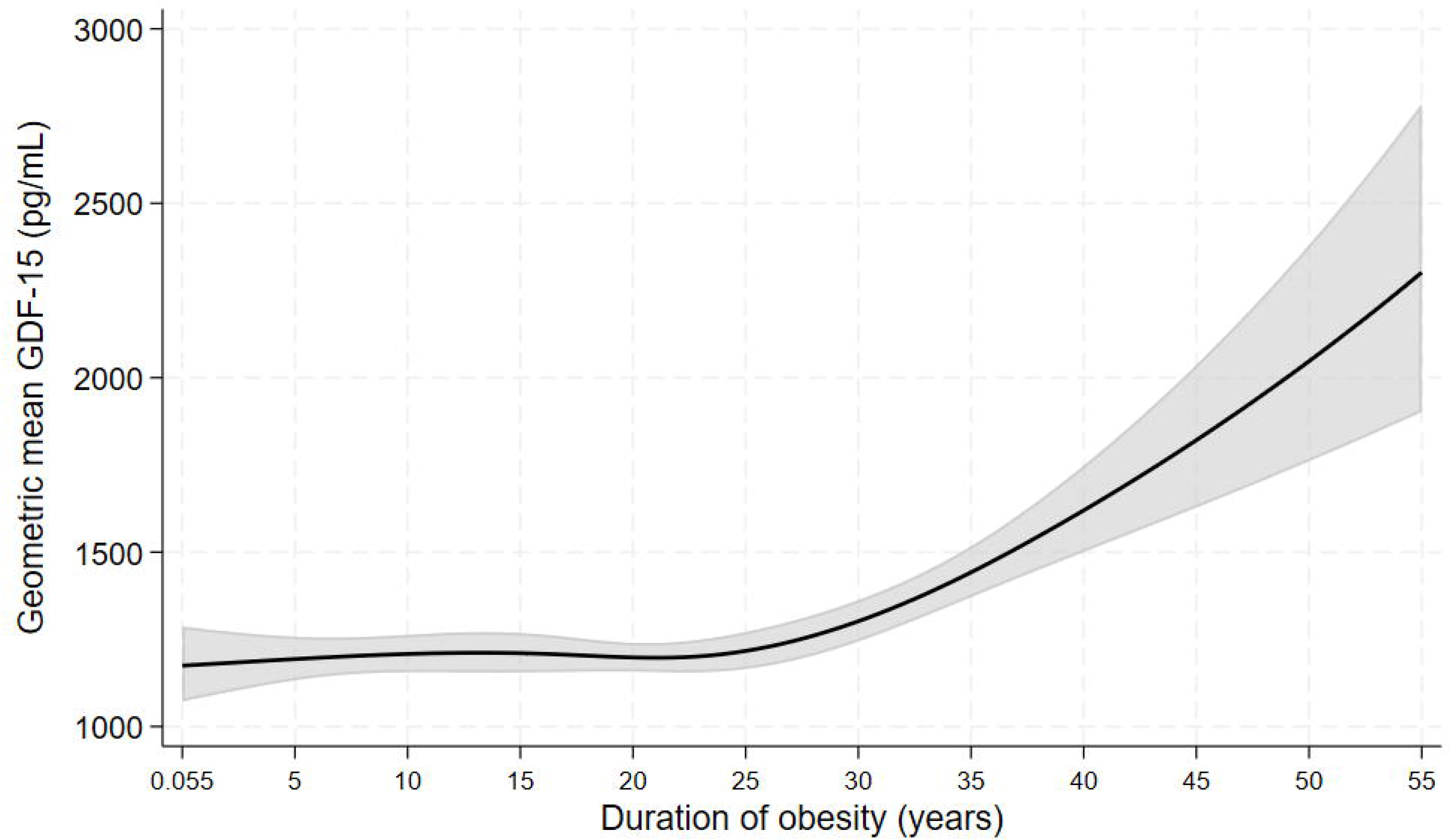
The association of obesity duration with GDF-15 (pg/mL) among participants who were ever obese, unadjusted for covariates in. **Table 1**

**Table 3.**
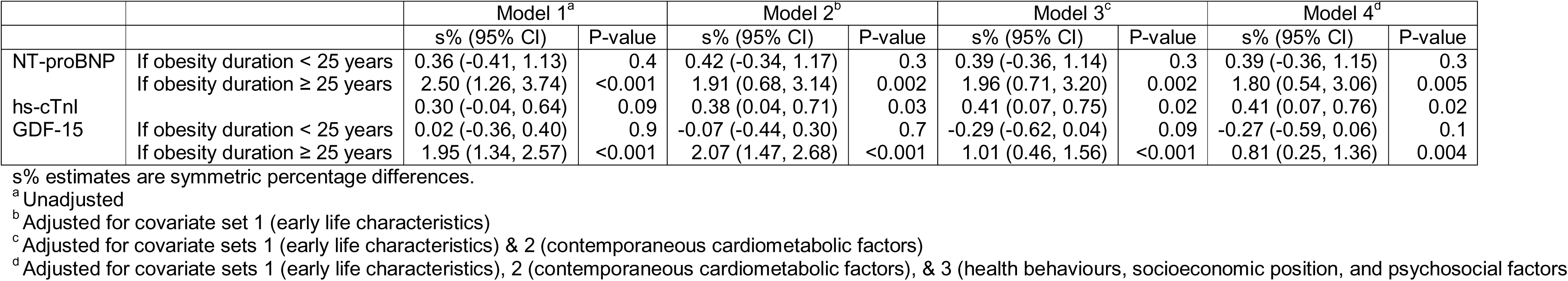
The associations of obesity duration (years) with NT-proBNP, hs-cTnI, and GDF-15 among participants who were ever obese.

|  |  | Model 1<sup>a</sup> |  | Model 2<sup>b</sup> |  | Model 3<sup>c</sup> |  | Model 4<sup>d</sup> |  |
| --- | --- | --- | --- | --- | --- | --- | --- | --- | --- |
|  |  | s% (95% CI) | P-value | s% (95% CI) | P-value | s% (95% CI) | P-value | s% (95% CI) | P-value |
| NT-proBNP | If obesity duration < 25 years | 0.36 (-0.41, 1.13) | 0.4 | 0.42 (-0.34, 1.17) | 0.3 | 0.39 (-0.36, 1.14) | 0.3 | 0.39 (-0.36, 1.15) | 0.3 |
|  | If obesity duration ≥ 25 years | 2.50 (1.26, 3.74) | <0.001 | 1.91 (0.68, 3.14) | 0.002 | 1.96 (0.71, 3.20) | 0.002 | 1.80 (0.54, 3.06) | 0.005 |
| hs-cTnI |  | 0.30 (-0.04, 0.64) | 0.09 | 0.38 (0.04, 0.71) | 0.03 | 0.41 (0.07, 0.75) | 0.02 | 0.41 (0.07, 0.76) | 0.02 |
| GDF-15 | If obesity duration < 25 years | 0.02 (-0.36, 0.40) | 0.9 | -0.07 (-0.44, 0.30) | 0.7 | -0.29 (-0.62, 0.04) | 0.09 | -0.27 (-0.59, 0.06) | 0.1 |
|  | If obesity duration ≥ 25 years | 1.95 (1.34, 2.57) | <0.001 | 2.07 (1.47, 2.68) | <0.001 | 1.01 (0.46, 1.56) | <0.001 | 0.81 (0.25, 1.36) | 0.004 |

s% estimates are symmetric percentage differences.
<sup>a</sup> Unadjusted
<sup>b</sup> Adjusted for covariate set 1 (early life characteristics)
<sup>c</sup> Adjusted for covariate sets 1 (early life characteristics) & 2 (contemporaneous cardiometabolic factors)
<sup>d</sup> Adjusted for covariate sets 1 (early life characteristics), 2 (contemporaneous cardiometabolic factors), & 3 (health behaviours, socioeconomic position, and psychosocial factors)

The associations of obesity duration with the three outcomes among participants who had ever experienced obesity, based on models unadjusted for covariates and models additionally adjusted for mean excess BMI (AUC/duration), are shown in Supplementary Figures 2–4. Visually, adjustment for mean excess BMI appeared to marginally attenuate the positive associations of obesity duration with both NT-proBNP and GDF-15 when duration exceeded 25 years. For hs-cTnI, the estimated positive association appeared slightly stronger after adjustment for mean excess BMI.

Supplementary Figures 5–7 show the associations between obesity duration and the three outcomes after adjustment for the different covariate sets. Based on visual inspection of the curves, the associations for NT-proBNP were broadly similar across the different levels of covariate adjustment. For hs-cTnI, the curves from the covariate-adjusted models were consistently higher than the unadjusted curve. Exploratory analyses in which covariates were added individually suggested that this difference was primarily attributable to adjustment for sex and General Ability Test score at age 11 years. For GDF-15, the curves from Models 3 and 4 appeared flatter beyond 25 years of obesity duration than those from Models 1 and 2. These observations were based on visual comparison of the curves, supplemented by exploratory analyses of individual covariate contributions, rather than formal statistical comparisons between curves.

Among participants who had ever experienced obesity, the associations of mean excess BMI with the three outcomes are shown in Supplementary Table 1. Mean excess BMI was positively associated with NT-proBNP across all models, with the association attenuating modestly after adjustment for early-life characteristics and remaining relatively stable with further adjustment. In Model 4, each 1 kg/m² higher mean excess BMI was associated with a 4.02% (95% CI 1.79–6.24) higher NT-proBNP. For hs-cTnI, there was no evidence of an association in the unadjusted model, but the estimate became larger and statistically significant after covariate adjustment. Exploratory analyses in which covariates were added individually suggested that this change was primarily attributable to adjustment for sex. In Model 4, each 1 kg/m² higher mean excess BMI was associated with a 2.00% (95% CI 0.49– 3.51) higher hs-cTnI. For GDF-15, mean excess BMI was positively associated with GDF-15 in Models 1 and 2, but there was no evidence of an association after adjustment for contemporaneous cardiometabolic factors in Models 3 and 4.

## Discussion

Understanding whether the duration and severity of obesity are associated with subclinical myocardial and cellular stress may help clarify the long-term cardiovascular consequences of obesity and provide context for life-course approaches to cardiovascular risk assessment. In this prospective birth cohort followed from childhood to later adulthood, longer duration of obesity was associated with higher concentrations of biomarkers reflecting myocardial stress, myocardial injury, and cellular stress at age 62 years. The associations differed across biomarkers: hs-cTnI increased approximately linearly with each additional year of obesity, whereas NT-proBNP and GDF-15 showed little evidence of association before approximately 25 years of obesity, after which concentrations increased progressively with duration. Among participants who had ever experienced obesity, greater mean excess BMI was associated with higher concentrations of NT-proBNP and hs-cTnI after full adjustment, whereas the association with GDF-15 attenuated substantially after adjustment for contemporaneous cardiometabolic factors. Taken together, these findings suggest that prolonged exposure to obesity may be particularly relevant to subclinical myocardial and cellular stress in later adulthood, with associations for NT-proBNP and GDF-15 becoming more evident after longer durations of obesity.

In cross-sectional studies, higher BMI and obesity have generally been associated with lower circulating NT-proBNP concentrations [10–12], which is a paradoxical finding given the greater haemodynamic load associated with obesity [42]. This inverse association may, however, reflect an obesity-related attenuation of the natriuretic peptide response to myocardial wall stress through mechanisms including reduced peptide synthesis or secretion and altered peptide clearance [43]. In contrast, we observed progressively higher NT-proBNP concentrations with longer duration of obesity, particularly after approximately 25 years. One possible explanation is that prolonged obesity-related haemodynamic load may contribute to progressive ventricular remodelling and subclinical myocardial dysfunction, which could result in greater myocardial wall stress with longer duration of obesity [44 45]. Our findings raise the possibility that the lower NT-proBNP concentrations observed in cross-sectional studies of individuals with obesity may not fully reflect the myocardial consequences of longstanding obesity.

In contrast to the pattern observed for NT-proBNP, hs-cTnI concentrations increased approximately linearly with longer duration of obesity, with each additional year of obesity associated with higher hs-cTnI concentrations after adjustment for early-life characteristics, contemporaneous cardiometabolic factors, health behaviours, socioeconomic position, and psychosocial factors. As hs-cTnI is a marker of myocardial injury, this finding suggests that prolonged exposure to obesity may be associated with greater subclinical myocardial injury in later adulthood. This association is consistent with cross-sectional evidence showing higher hs-cTnI concentrations among individuals with higher BMI in the general population [14]. We also observed higher hs-cTnI concentrations among men who had ever experienced obesity compared with men who had never experienced obesity, whereas there was no evidence of an association among women. Previous evidence indicates that the association between BMI and hs-cTnI may differ by sex, with the association between BMI and hs-cTnI being stronger in men than women [14 46]. For example, in the longitudinal Whitehall II study, which used repeated measures of BMI and hs-cTnI, each 5 kg/m² higher BMI was associated with 6% higher hs-cTnI concentrations among men (ratio, 1.06; 95% CI, 1.04–1.08), whereas there was no evidence of an association among women (ratio, 1.01; 95% CI, 0.99–1.04; P_interaction=0.008) [46]. This sex difference may partly reflect differences in body-fat distribution, with men generally having a greater proportion of visceral or abdominal adipose tissue, as well as sex differences in cardiac structure and the effects of sex hormones on cardiovascular physiology [46 47].

Among participants who had ever experienced obesity, greater mean excess BMI was associated with higher GDF-15 concentrations in models adjusted for early-life characteristics, but this association was substantially attenuated after adjustment for contemporaneous cardiometabolic factors and was no longer evident after full adjustment. This attenuation is consistent with evidence that GDF-15 is closely associated with cardiometabolic dysfunction. In obese individuals, GDF-15 has been associated with insulin resistance and several measures of glucose metabolism, including insulin, glucose, and HbA1c [48]. In the Atherosclerosis Risk in Communities Study, higher GDF-15 concentrations were associated with diabetes, atherosclerotic cardiovascular disease, heart failure, and markers of subclinical myocardial stress and injury [49]. More broadly, GDF-15 has been characterised as a stress-responsive cytokine associated with obesity, insulin resistance, diabetes, and cardiovascular disease [50]. Together, these findings suggest that the relationship between obesity and GDF-15 may reflect broader metabolic and cardiovascular stress rather than adiposity alone.

Taken together, the findings across the three biomarkers suggest that longer exposure to obesity may be associated with greater subclinical myocardial and cellular stress in later adulthood. Importantly, the associations between obesity duration and all three biomarkers persisted (albeit with some attenuation for NT-proBNP and GDF-15) after adjustment for mean excess BMI, indicating that the observed relationships were not simply a reflection of greater cumulative severity among those with longer exposure to obesity. Participants with longer durations of obesity had higher biomarker concentrations even after accounting for the extent to which their BMI exceeded the age-specific obesity threshold. This suggests that the length of time spent with obesity may capture an aspect of cumulative obesity exposure that is distinct from the magnitude of excess adiposity. Obesity duration was strongly inversely correlated with age at onset (Spearman’s ρ=−0.96), and these findings are therefore also consistent with an association between earlier onset of obesity and higher biomarker concentrations at age 62 years.

From a clinical and public health perspective, our findings underscore the potential importance of the duration of obesity for later-life cardiovascular health. Current approaches to cardiovascular risk assessment primarily use established risk factors measured in adulthood, including age, blood pressure, smoking, and blood lipids [51 52]. Our findings suggest that the cumulative history of obesity across the life course may also be relevant to subclinical myocardial stress and injury. These findings provide further rationale for preventing the development of obesity early in the life course and supporting effective, sustained management among those who develop obesity, given the potential for prolonged exposure to obesity to affect cardiovascular health. Previous studies have incorporated the duration and severity of obesity into cumulative exposure measures and reported better model fit for cardiovascular disease than models based on BMI or obesity duration alone [15 18]. Whether obesity duration or age at onset provides incremental predictive value beyond established cardiovascular risk factors and current adiposity remains an important question for future research.

The key strengths of the present study are the prospective assessment of obesity from 7 to 62 years of age in a relatively large sample, with repeated BMI measurements used to model individual obesity trajectories and derive measures of obesity duration and cumulative severity, together with the availability of three complementary biomarkers of myocardial stress, myocardial injury, and cellular stress at age 62 years. Previous studies have examined the duration and severity of obesity across the life course in relation to cardiometabolic risk factors (e.g., blood pressure, HDL cholesterol, and HbA1c) or obesity duration during adulthood in relation to subclinical myocardial injury [17 22]. However, to our knowledge, no previous study has characterised obesity trajectories from childhood into late adulthood and examined their duration and severity in relation to biomarkers of subclinical myocardial stress, myocardial injury, and cellular stress measured in later life.

Several limitations should be considered. First, the analytic sample represented a subset of the original 1958 NCDS, reflecting attrition over follow-up and the availability of biomarker data. Participants included in the analysis may therefore differ systematically from those not included, potentially introducing selection bias, including collider bias if study participation is influenced by factors related to both the exposure and the outcome [53 54]. Second, although obesity was assessed prospectively from 7 to 62 years of age, biomarkers were measured only at age 62 years, precluding examination of their trajectories over time. Third, obesity was defined using BMI, which does not distinguish between adipose and lean mass or capture variation in adipose tissue distribution [55]. In addition, although measured and self-reported BMI observations were incorporated into the trajectory model with different residual variances, some BMI measurements were self-reported and may therefore be subject to measurement error. BMI trajectories were also modelled from measurements obtained at discrete ages, and therefore the derived duration and cumulative severity measures are estimates rather than direct observations of continuous obesity exposure. Fourth, obesity duration was strongly inversely correlated with age at onset (Spearman’s ρ=−0.96), making it difficult to distinguish associations of longer duration from those of earlier onset of obesity. Finally, although we adjusted for a wide range of early-life and adult characteristics, residual confounding cannot be excluded given the observational nature of the study. Some of the adult covariates may also lie on the causal pathway between prolonged obesity and biomarker concentrations, and adjustment for such variables could therefore have introduced bias by controlling for potential mediators [56]. The cohort was also born in 1958 and was exposed to an obesity and cardiovascular risk environment that differs from that of more recent generations, which may limit the generalisability of the findings to contemporary populations.

To conclude, in this prospective birth cohort, longer duration of obesity from childhood to later adulthood was associated with higher concentrations of biomarkers of myocardial stress and injury at age 62 years, with associations for NT-proBNP and GDF-15 becoming more evident after longer durations, whereas hs-cTnI increased across the duration range. Greater cumulative severity of obesity was also associated with higher concentrations of NT-proBNP and hs-cTnI. These findings indicate that the cardiovascular consequences of obesity may reflect not only the severity of excess adiposity but also the length of time individuals are exposed to obesity. Consideration of cumulative obesity exposure across the life course may therefore be relevant to understanding subclinical cardiovascular injury and stress in later adulthood.

## Data Availability

All data used in the present study are freely available to download via the UK Data Service (https://ukdataservice.ac.uk/).

## Acknowledgments

We gratefully acknowledge the 1958 NCDS cohort members whose longstanding participation has made this research possible. Their continued engagement with the study has provided an invaluable resource for life-course research and will support important research in the years to come.

## Funding

This paper received no direct funding. WJ acknowledges support from the National Institute for Health and Care Research (NIHR) Leicester Biomedical Research Centre, which is a partnership between University Hospitals of Leicester NHS Trust, Loughborough University, and the University of Leicester. The views expressed are those of the authors and not necessarily those of the NHS, the NIHR, or the Department of Health. GDB is supported by the UK Medical Research Council (MR/P023444/1) and the US National Institute on Aging (1R56AG052519-01, 1R01AG052519-01A1).

Disclosures

All authors declare that they have no conflict of interest. TN is a full-time employee of AstraZeneca outside of this research.

Author contributions

WJ and DB conceived and developed the research idea. WJ and TN conducted the statistical analyses. WJ drafted the manuscript. DB and MH secured funding for the study. All authors contributed to the interpretation of the findings, critically revised the manuscript for important intellectual content, and approved the final version.

