## Supplementary Material for "Impact of Prolonged Obesity Across the Lifecourse on Myocardial Stress and Injury in Older Age: Birth Cohort Study"

Supplementary Figure 1. Histogram of duration of obesity

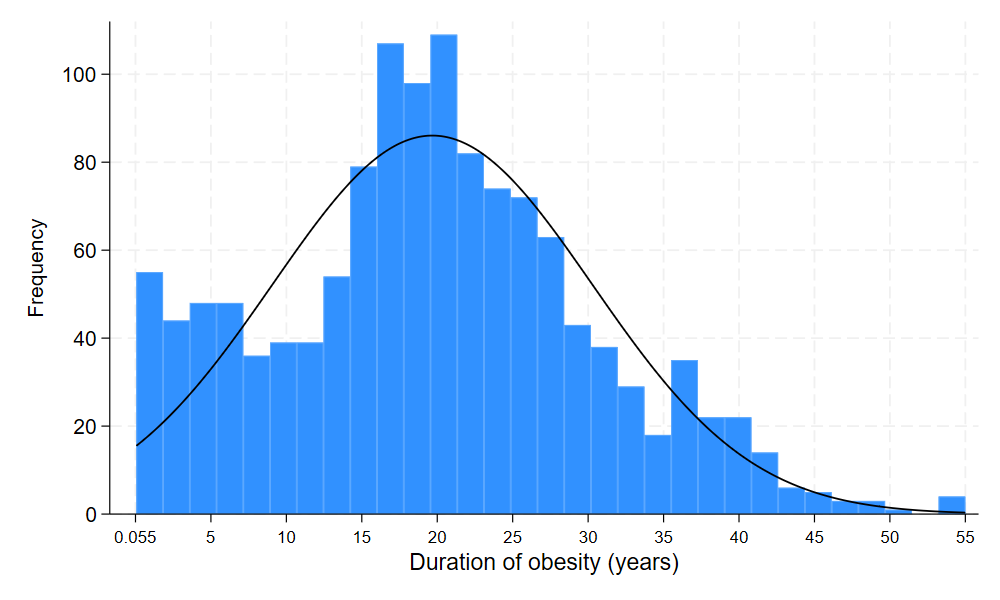

Supplementary Figure 2. The association of obesity duration with NT-proBNP (pg/mL) among participants who were ever obese, before and after adjustment for mean excess BMI (AUC/duration)

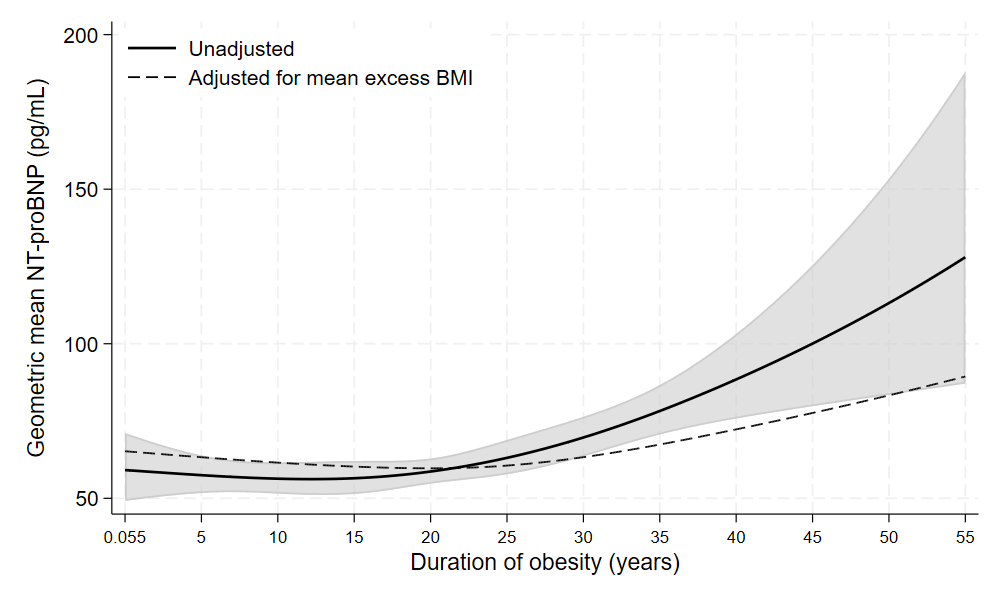

Supplementary Figure 3. The association of obesity duration with hs-cTnI (pg/mL) among participants who were ever obese, before and after adjustment for mean excess BMI (AUC/duration)

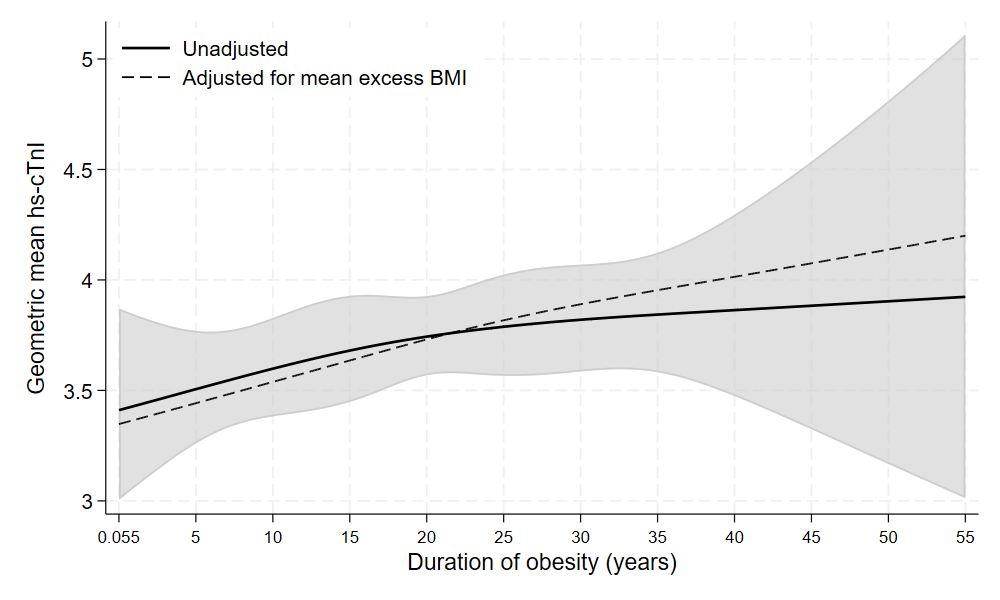

Supplementary Figure 4. The association of obesity duration with GDF-15 (pg/mL) among participants who were ever obese, before and after adjustment for mean excess BMI (AUC/duration)

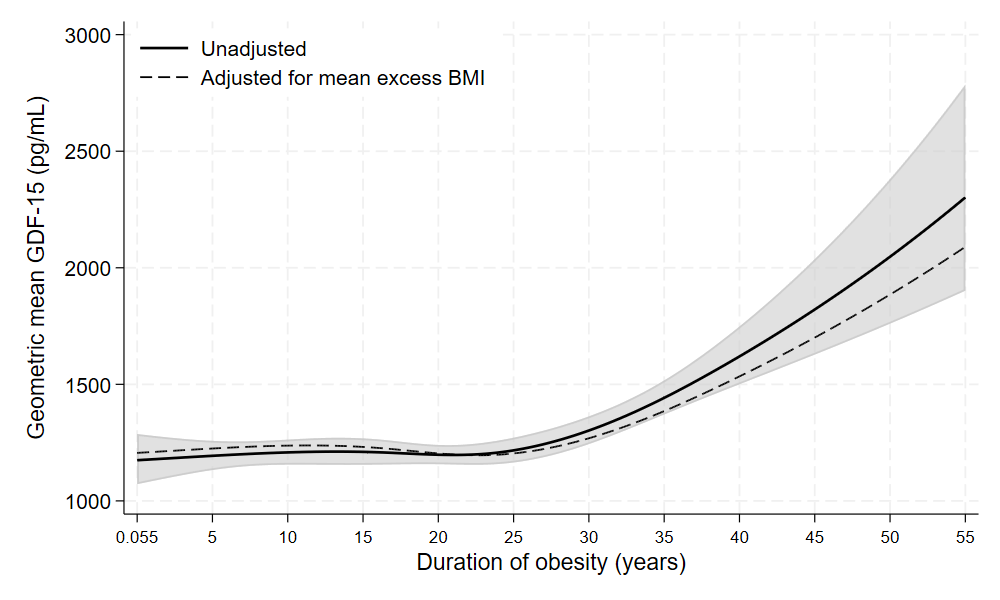

Supplementary Figure 5. The association of obesity duration with NT-proBNP (pg/mL) among participants who were ever obese, before and after adjustment for different covariates sets

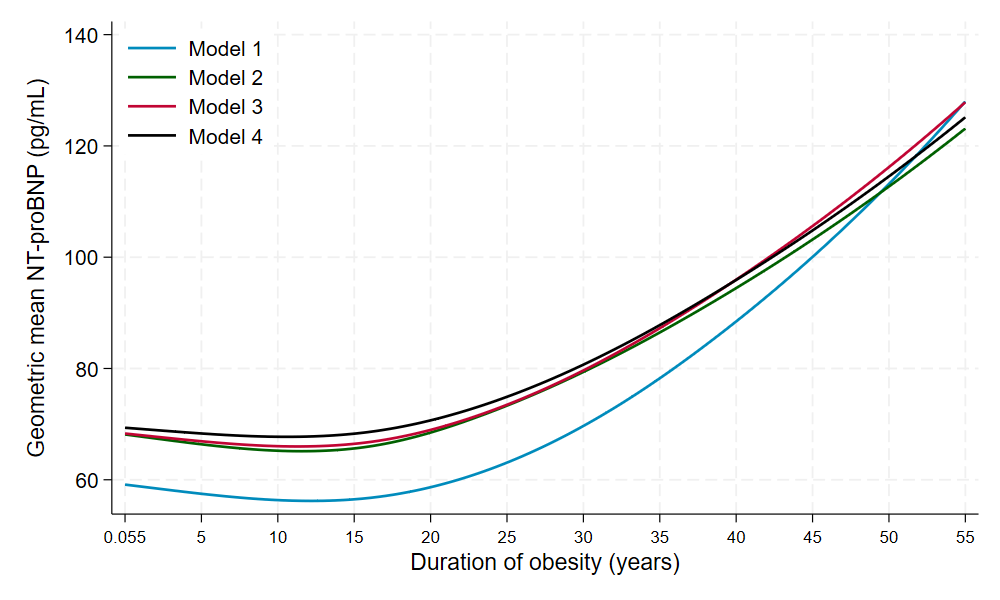

Model 1: Unadjusted

Model 2: Adjusted for covariate set 1 (early life characteristics)

Model 3: Adjusted for covariate sets 1 (early life characteristics) & 2 (contemporaneous cardiometabolic factors)

Model 4: Adjusted for covariate sets 1 (early life characteristics), 2 (contemporaneous cardiometabolic factors), & 3 (health behaviours, socioeconomic position, and psychosocial factors

Supplementary Figure 6. The association of obesity duration with hs-cTnI (pg/mL) among participants who were ever obese, before and after adjustment for different covariates sets

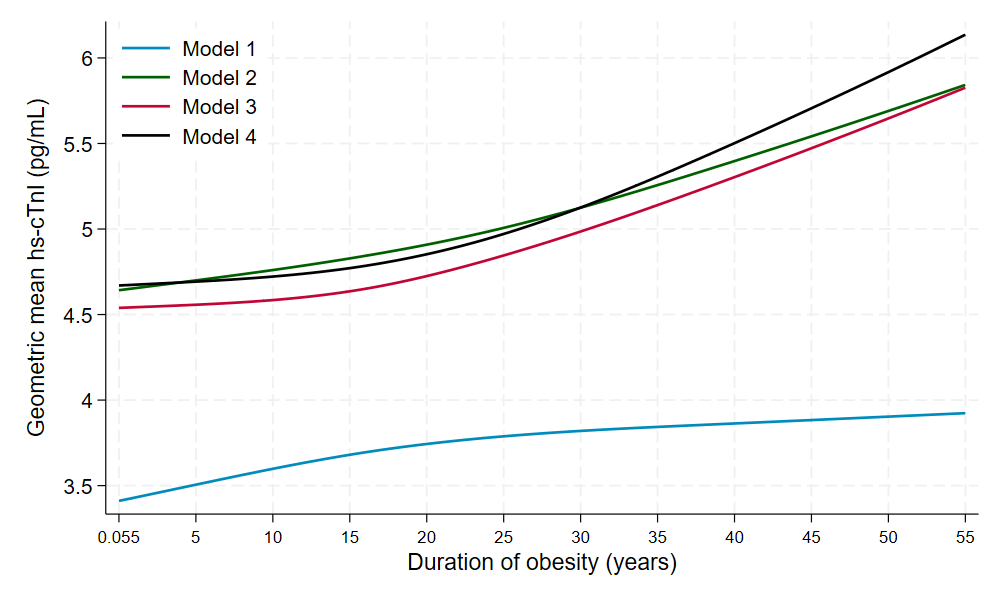

Model 1: Unadjusted

Model 2: Adjusted for covariate set 1 (early life characteristics)

Model 3: Adjusted for covariate sets 1 (early life characteristics) & 2 (contemporaneous cardiometabolic factors)

Model 4: Adjusted for covariate sets 1 (early life characteristics), 2 (contemporaneous cardiometabolic factors), & 3 (health behaviours, socioeconomic position, and psychosocial factors

Supplementary Figure 7. The association of obesity duration with GDF-15 (pg/mL) among participants who were ever obese, before and after adjustment for different covariates sets

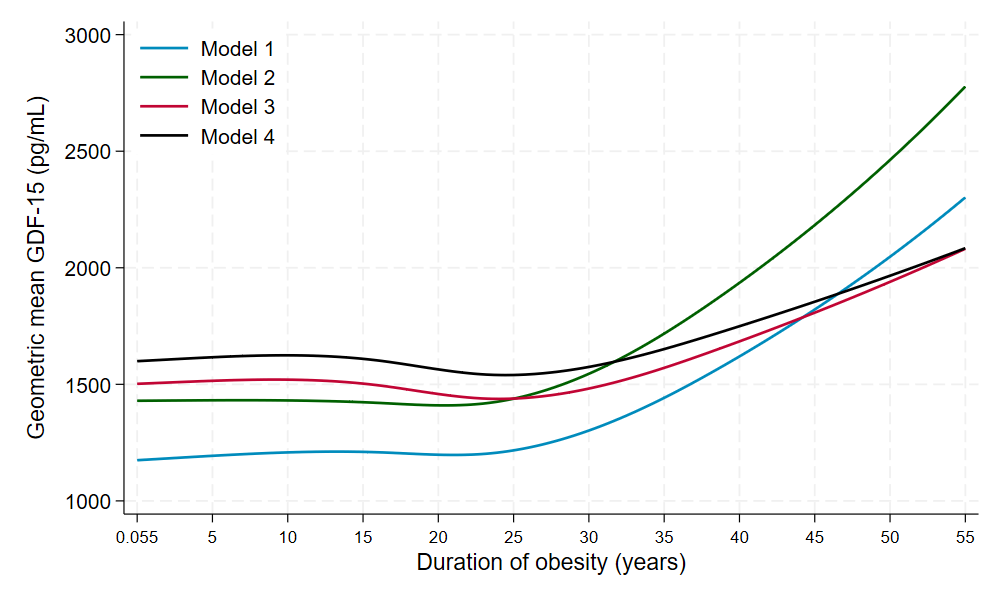

Model 1: Unadjusted

Model 2: Adjusted for covariate set 1 (early life characteristics)

Model 3: Adjusted for covariate sets 1 (early life characteristics) & 2 (contemporaneous cardiometabolic factors)

Model 4: Adjusted for covariate sets 1 (early life characteristics), 2 (contemporaneous cardiometabolic factors), & 3 (health behaviours, socioeconomic position, and psychosocial factors

Supplementary Table 1. The associations of mean excess BMI (AUC/duration) with NT-proBNP, hs-cTnI, and GDF-15 among participants who were ever obese

|  | Model 1^a^ | | Model 2^b^ | | Model 3^c^ | | Model 4^d^ | |
| --- | --- | --- | --- | --- | --- | --- | --- | --- |
|  | s% (95% CI) | P-value | s% (95% CI) | P-value | s% (95% CI) | P-value | s% (95% CI) | P-value |
| NT-proBNP | 5.74 (3.67, 7.82) | <0.001 | 4.19 (2.10, 6.28) | <0.001 | 4.26 (2.10, 6.43) | <0.001 | 4.02 (1.79, 6.24) | <0.001 |
| hs-cTnI | 0.58 (-0.85, 2.02) | 0.4 | 1.75 (0.33, 3.18) | 0.02 | 1.90 (0.44, 3.37) | 0.01 | 2.00 (0.49, 3.51) | 0.009 |
| GDF-15 | 2.99 (4.03, 0.00) | <0.001 | 3.29 (4.32, 0.00) | <0.001 | 0.79 (1.75, 0.00) | 0.1 | 0.51 (1.48, 0.00) | 0.3 |

s% estimates are symmetric percentage differences.

^a^ Unadjusted

^b^ Adjusted for covariate set 1 (early life characteristics)

^c^ Adjusted for covariate sets 1 (early life characteristics) & 2 (contemporaneous cardiometabolic factors)

^d^ Adjusted for covariate sets 1 (early life characteristics), 2 (contemporaneous cardiometabolic factors), & 3 (health behaviours, socioeconomic position, and psychosocial factors
